# The changing characteristics of COVID-19 presentations: A regional comparison of SARS-CoV-2 hospitalised patients during the first and second wave

**DOI:** 10.1101/2021.02.07.21251297

**Authors:** Catherine Atkin, Vicky Kamwa, V Reddy-Kolanu, D Parekh, Felicity Evison, Peter Nightingale, Suzy Gallier, Simon Ball, Elizabeth Sapey

## Abstract

**Background:** This study assesses COVID-19 hospitalised patient demography and outcomes during wave 1 and wave 2, prior to new variants of the virus.

**Methods:** All patients with a positive SARS-CoV-2 swab between 10^th^ March 2020 and 5^th^ July 2020 (wave 1) and 1^st^ September 2020 and 16^th^ November 2020 (wave 2) admitted to University Hospitals Birmingham NHS Foundation Trust were included (n=4856), followed for 28 days.

**Results:** Wave 2 patients were younger, more ethnically diverse, had less co-morbidities and disease presentation was milder on presentation. After matching for these factors, mortality was reduced, but without differences in intensive care admissions.

**Conclusion:** Prior to new SARS-CoV-2 variants, outcomes for hospitalised patients with COVID-19 were improving but with similar intensive care needs.

## Introduction

The global pandemic caused by SARS-CoV-2 continues to provide significant health challenges worldwide, with a high number of admissions to hospital and a high mortality rate. The prevalence and impact of infection is heterogeneous in individual countries and in different regions within countries(1). In the UK, there have been waves of COVID-19-associated hospitalisations. Outcomes for patients infected with SARS-CoV-2 can be altered by the characteristics of people exposed to the virus (which can be influenced by social restrictions), medical interventions and variations in the virus itself, including new mutations which might affect infectivity or virulence.

The UK has implemented a number of social restriction measures. In England, there have been three national lockdowns (26^th^ March 2020; 5^th^ November 2020 and 4^th^ January 2021 (2)), intermittent shielding for the elderly or those with significant health conditions and regional restrictions leading to a tier system(3). The allocation of a tier in the UK reflects local infection rates and hospital admissions/ bed capacity. The viral caseload after the first lockdown was significantly reduced and the most severe social restrictions eased initially on 4th July 2020(4). Face-coverings were mandated in shops and supermarkets from 24^th^ July 2020(5). A patchwork of local restrictions were implemented from September 2020 and the tier system was first introduced on 14th October 2020(6).

These measures are designed to limit viral spread, protect the most vulnerable and ensure local health services are not overwhelmed. In practice, these evolving social measures alter the demographics of people most likely to be exposed to the virus. However, the natural history of SARS-CoV-2 infectivity means that there is a delay between the implementation of social restrictions, a subsequent reduction in infection rates, and then a reduction in hospitalisation and death.

Scientific discovery has led to improvements in patient care during the pandemic. Clinical trials have reported beneficial or ineffective treatments in specific care settings or patient groups (7, 8). Emerging guidelines have provided standardised care pathways(9) and supported anti-microbial stewardship(10).

As well as social policy and scientific advancements, the prevalence and outcomes of COVID-19 could be affected by changes to the virus. SARS-CoV-2 variants have been described in most countries(11). “Variants under investigation” are declared when specific changes to the virus may alter its impact on human health. The VUI-202012/01 (the first “Variant Under Investigation”) has 17 mutations including an N501Y mutation in the spike protein that the virus uses to bind to the human ACE2 receptor. Changes in this part of spike protein have been associated with increased infectivity and spread(12) and this variant has been linked with increased cases of COVID-19, initially in the south east of the UK (13).

Combined datasets, such as the global ISARIC study, provide an analysis of the overall spectrum of disease and average impact on health services(14). However, to understand the evolving impact of COVID-19, one must account for the differential regional spread of the virus, differences in regional social restrictions and the adoption of new treatment pathways, before one can assess the significance of new variants of the virus, which have variable penetration across regions of the UK and internationally.

Birmingham is one of the most ethnically diverse cities in the UK with a high burden of COVID-19 in all the UK COVID-19 waves. Following a rise in cases in the early autumn, Birmingham was placed under a high level of social restriction from 15^th^ September 2020 which lasted until the third national lockdown. Up until mid-November 2020, the prevalence of the VUI-202012/01 variant among Birmingham cases was low(13).

This study was conducted to understand changes in hospitalised COVID comparing a period before and after social restrictions were in place and new treatments were available, and before the new variant was dominant in Birmingham. We aimed to assess whether the population of patients admitted to hospital had changed during evolving health policy decisions, whether this impacted on the severity of illness and outcomes and whether new treatments and guidance (specifically antibiotic stewardship) had been adopted.

## Methods

This data study was supported by PIONEER, a Health Data Research Hub in Acute Care with ethical approval provided by the East Midlands – Derby REC (reference: 20/EM/0158).

University Hospitals Birmingham NHS Foundation Trust (UHB), UK is one of the largest Trusts nationally, covering 4 NHS hospital sites, treating over 2.2 million patients per year and housing the largest single critical care unit (CCU) in Europe. UHB saw the highest number of COVID admissions in the UK (6047 confirmed cases by 29th December 2020) and the highest number of patients ventilated, with an expanded CCU capacity of >200 beds.

### Study population

All patients with a confirmed positive severe acute respiratory syndrome coronavirus-2 (SARS-CoV-2) swab result between 10^th^ March 2020 and 5^th^ July 2020 and from 1^st^ September 2020 and 16^th^ November 2020 taken up to 14 days before and 7 days after admission to UHB were included. COVID cases were confirmed following a nasopharyngeal and oropharyngeal swab in all cases(15) which were processed in accordance with NHS guidance within UHB NHS laboratories (16).

Dates were chosen to include the first confirmed SARS-CoV-2 case in UHB. The end date for the first wave was chosen as confirmed SARS-CoV-2 positive admissions fell to a low plateau between 5^th^ July 2020 and 1^st^ September. Admissions started to rise from 1^st^ September 2020, which was chosen as the start date for the second period. Admissions associated with positive swabs taken after 16^th^ November were not included, to minimise the inclusion of the new variant (designated VUI-202012/01 (the first Variant Under Investigation in December 2020)(17)), as this variant may have altered virulence as well as infectivity.

UHB has built and runs its own electronic health record (EHR) and was able to develop a structured electronic clerking proforma where all patients suspected of having COVID-19 could be identified on admission. In Wave 2, all admitted patients were swabbed for SARS-CoV-2 irrespective of suspicion of COVID, on admission, during the admission and on discharge.

For all patients, the results of the first positive swab were included but patient records were checked for subsequent positive swab results if associated with a subsequent admission. Mortality and (in those alive) patient admission status (discharged and alive, continued admission and alive) were assessed 28 days after the first positive swab result (the latest date to assess outcome being 14^th^ December 2020).

### Data Collection and variable definitions

Patient demographics and clinical data were collected from the UHB EHR. Clinician confirmed co-morbidities were available from the EHR, the summary primary care record (Your Care Connected) and from diagnostic codes derived from previous hospital episodes. The EHR encodes diagnoses using NHS Digital SNOMED CT browser(18) alongside and mapped on to ICD-10 codes(19) allowing inclusion of historically entered ICD10 codes. A simple count of co-morbidities was undertaken to determine the impact of multi-morbidity, as described (20).

English Indices of Deprivation scores were calculated using postcodes from the current data provided by the UK’s Ministry of Housing, Communities and Local Government (2019) Report(21). Ethnicity was self-reported by the patient or their family members on admission to hospital. Where this data was missing, it was gathered from previous admissions and by reviewing primary and secondary medical records. Ethnicity was grouped as per national guidelines(22) and was classified as unknown in <6% of patients.

### Severity of COVID-19 on admission

The physician determined severity of COVID-19 on first admission was categorised using a pragmatic and locally developed score to identify those on admission to hospital who were in need of urgent critical care assessments for respiratory support, as previously described(23) and is as follows:

1. Patients were considered to have severe respiratory manifestations of COVID-19 infection; if COVID was suspected and the patient required inspired oxygen ≥ 50% to maintain targeted oxygen saturations (>93% except in the presence of type 2 respiratory failure where the target saturations were 88% - 92%) with respiratory pathology thought driven by COVID-19 illness.
2. If not severe, patients were considered to have moderate severity respiratory manifestations of COVID-19 infection if COVID was suspected and the patient required inspired oxygen of > 4L/min or inspired oxygen > 28% to maintain target oxygen saturations.
3. Patients were considered to have mild severity respiratory manifestations of COVID-19 infection if the patient had respiratory symptoms but did not meet the severe or moderate criteria as described above.

Baseline physiological assessments to determine severity of COVID-19 were considered to be those taken within 24 hours either side of the SARS-CoV-2 swab collection time, of which the earliest available measurement was used. As not all patients were admitted within 24 hours of their SARS-CoV-2 swab test, and since these assessments are only routinely recorded in the EHR system for Queen Elizabeth Hospital Birmingham patients, baseline severity scores were only available for a subset of the patients (3363/4856).

To determine if disease severity on admission reflected duration of illness, medical clerking notes were reviewed to determine the duration of symptoms prior to admission. This was available in only a subset of patients (1619/4856). COVID-associated prescribing and administration (dexamethasone, remdesivir and broad-spectrum antibiotics or those used for respiratory tract infections) were collated from the EHR at UHB. Drugs were included where drug prescription and administration continued after the confirmed SARS-CoV-2 swab results were known and reviewed by a clinician.

### Outcomes

The primary outcome was death while in hospital or post discharge within 28 days of a positive SARS-CoV-2 test, as per national reporting(24). For those patients discharged from hospital, primary care records were checked and any patients admitted to hospital with COVID-19 and discharged who had died in the community within the censor period were noted. Those with an on-going admission were censored 28 days after a positive swab result.

### Statistics

Statistical analysis was performed using STATA (SE) version 15. Baseline characteristics for the total population and ethnic communities are presented as mean (standard deviation) or median (interquartile range) for continuous variables and as frequency (percentage) for categorical variables. Continuous variables were compared between data sets using Mann-Whitney U tests. Categorical variables were compared using Fisher exact and Chi-Square tests. We assessed whether differences in mortality from wave 1 to wave 2 could be accounted for differences in the population by matching patients by age, ethnic group and co-morbidity counts. To improve the balance between the waves, coarsened exact matching was performed using R statistical software (4.03) and using the package MatchIt. Balance was compared before and after matching. Results were considered significant if the p-value was <0.05.

## Results

In total, 4856 confirmed cases were assessed. There were 2949 SARS-CoV-2 swab positive patients admitted during Wave 1 and 1907 admitted during the included dates for Wave 2. Figure 1a shows all COVID swab positive admissions between 10^th^ March to 16^th^ November 2020 plotted as days since start of wave when the swab was taken.

**Figure 1a.**
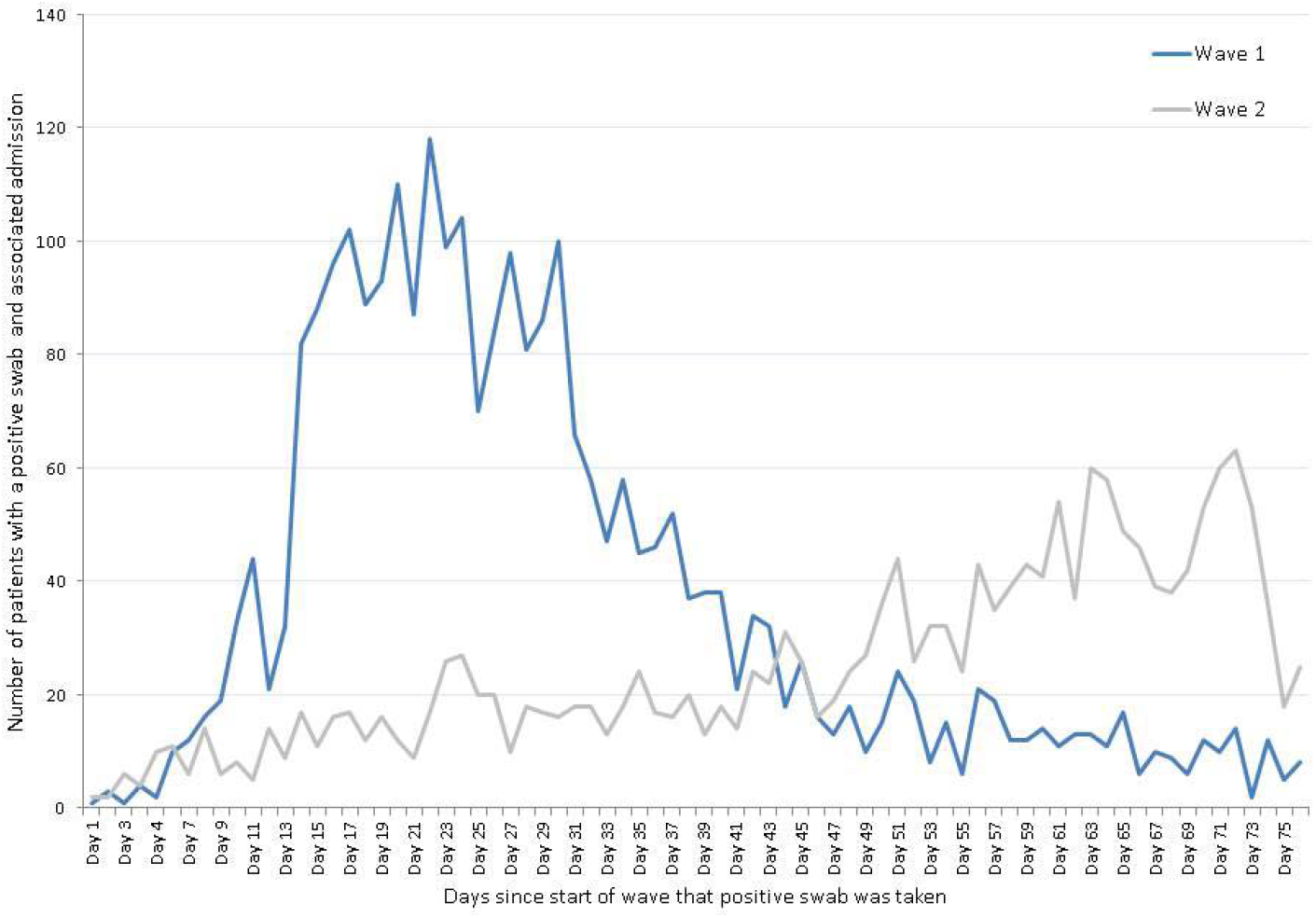
Number of patients with a positive swab and associated admission to hospital. **Legend**. The number of patients for wave 1 and wave 2 who had a positive swab and associated admission plotted against the days since the start of the wave. These days are within the dates as given.

**Figure 1b.**
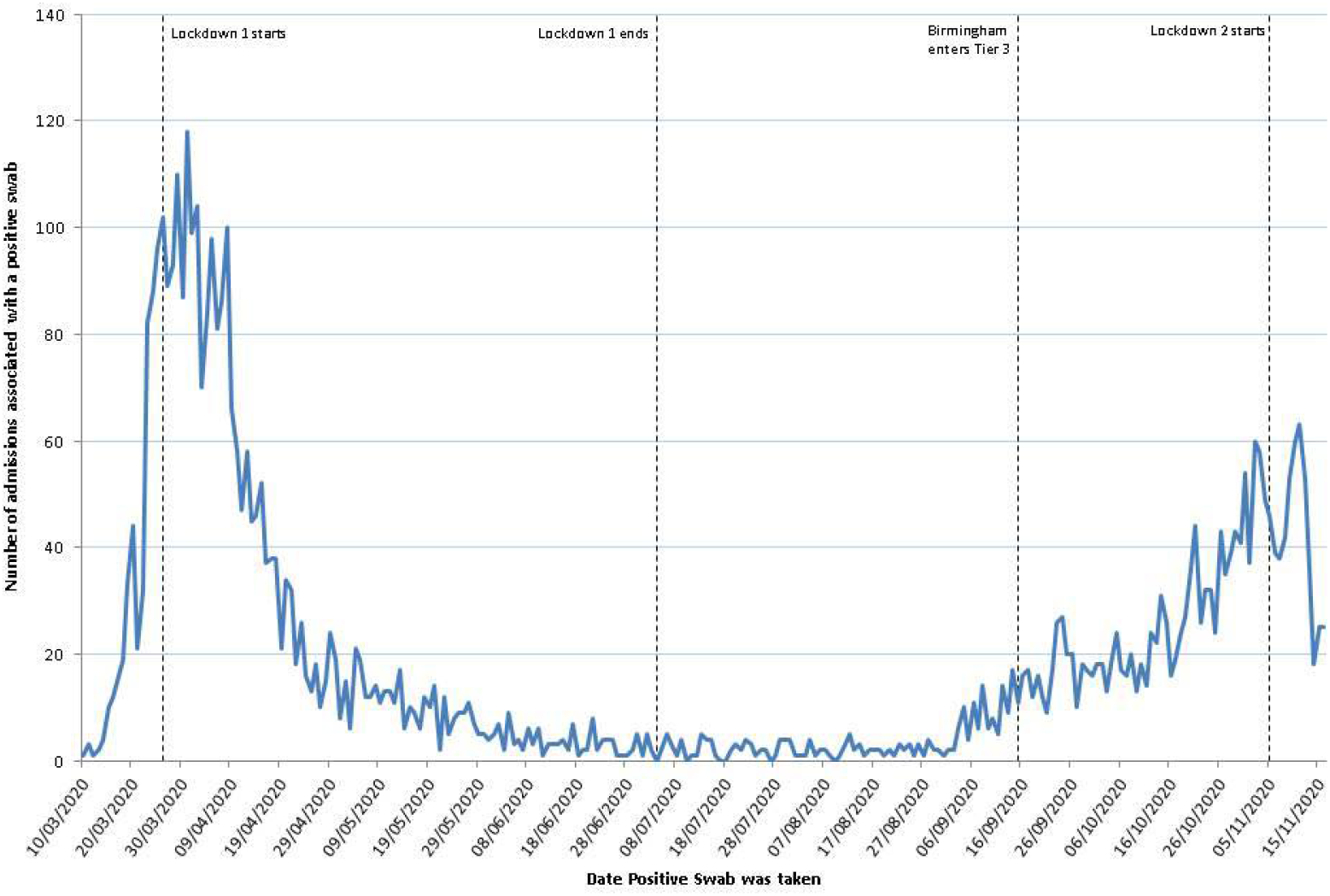
Number of patients with a positive swab and associated admission to hospital. The number of patients per day who had a positive swab and associated admission plotted against the date the swab was taken, this shows data for the whole of the time period, including “between waves”.

Four of the included patients (0.08%) had a positive COVID test in wave 1 and a subsequent positive COVID test in wave 2. No patients had received a SARS-CoV-2 vaccine in this population.

### Patient demographics

In comparison to wave 1, admitted wave 2 patients were younger, less likely to be of white ethnicity, and lived in areas associated with more social deprivation. They were also less likely to have co-morbid disease than in Wave 1, with a median co-morbidity count of 3 (IQR 1 -4) (wave 1) versus 2 (IQR 0-4) (wave 2), p<0.001. There was no difference in median BMI comparing Wave 1 (27 (IQR 24 – 33)) to wave 2 (28 (IQR 25 – 33)), p = 0.0503, with 70.0% of all admitted patients with a BMI reading measured, being overweight, obese or morbidly obese. The demographics of included patients are shown in Table 1.

**Table 1.** A comparison of demographics between patients admitted 10^th^ March – 5^th^ July 2020 and 1^st^ September to 3^rd^ November 2020.

|  |  | Wave 1 | Wave 2 | p-value |
| --- | --- | --- | --- | --- |
| Overall Count |  | 2949 | 1907 |  |
| Sex | Female | 1346 (45.6%) | 880 (46.1%) | p=0.731 |
|  | Male | 1603 (54.4%) | 1027 (53.9%) |  |
| Age | Median (IQR) | 73 (59-84) | 68 (51-81) | p<0.001 |
|  | 50 and Under | 474 (16.1%) | 454 (23.8%) | p<0.001 |
|  | 51-65 | 555 (18.8%) | 421 (22.1%) |  |
|  | 66-75 | 558 (18.9%) | 321 (16.8%) |  |
|  | 76-85 | 748 (25.4%) | 440 (23.1%) |  |
|  | Over 85 | 614 (20.8%) | 271 (14.2%) |  |
| Ethnicity | White | 2063 (70.0%) | 1131 (59.3%) | p<0.001 |
|  | Black | 172 (5.8%) | 89 (4.7%) |  |
|  | South Asian | 475 (16.1%) | 497 (26.1%) |  |
|  | Mixed | 28 (0.9%) | 22 (1.2%) |  |
|  | Chinese | 20 (0.7%) | 4 (0.2%) |  |
|  | Any Other Ethnic Group | 67 (2.3%) | 39 (2.0%) |  |
|  | Unknown | 124 (4.2%) | 125 (6.6%) |  |
| IMD Quintile | 1 (Most Deprived) | 1340 (45.4%) | 1035 (54.3%) | p<0.001 |
|  | 2 | 570 (19.3%) | 295 (15.5%) |  |
|  | 3 | 437 (14.8%) | 250 (13.1%) |  |
|  | 4 | 277 (9.4%) | 169 (8.9%) |  |
|  | 5 (Least Deprived) | 310 (10.5%) | 152 (8.0%) |  |
|  | Unknown | 15 (0.5%) | 6 (0.3%) |  |
| Co-morbidities* | Chronic Kidney Disease | 784 (26.6%) | 363 (19.0%) | p<0.001 |
|  | Dementia | 589 (20.0%) | 195 (10.2%) | p<0.001 |
|  | Interstitial Lung Disease | 113 (3.8%) | 55 (2.9%) | p=0.078 |
|  | Stroke or cerebrovascular | 774 (26.2%) | 319 (16.7%) | p<0.001 |
|  | Ischaemic heart disease | 908 (30.8%) | 495 (26.0%) | p<0.001 |
|  | Asthma | 572 (19.4%) | 355 (18.6%) | p=0.499 |
|  | Hypertension | 2030 (68.8%) | 1018 (53.4%) | p<0.001 |
|  | Diabetes | 1041 (35.3%) | 590 (30.9%) | p=0.002 |
|  | Any active malignancy | 596 (20.2%) | 260 (13.6%) | p<0.001 |
|  | COPD | 579 (19.6%) | 272 (14.3%) | p<0.001 |
|  | Atrial fibrillation | 807 (27.4%) | 336 (17.6%) | p<0.001 |
| BMI (n = 1420) | Underweight | 30 (3.2%) | 11 (2.3%) | p = 0.05 |
|  | Normal weight | 264 (28.2%) | 122 (25.3%) |  |
|  | Overweight | 307 (32.8%) | 153 (31.7%) |  |
|  | Obese | 279 (29.8%) | 157 (32.5%) |  |
|  | Morbid obesity | 57 (6.1%) | 40 (8.3%) |  |

**Legend:** Data is number (percentage) unless otherwise stated. Ethnicity was self-reported (see Methods). English Indices of deprivation (IMD) were calculated using postcode. Diabetes includes type 1 and type 2 diabetes. COPD= Chronic Obstructive Pulmonary Disease. Patients could (and often did) have more than one co-morbid condition

### Disease presentation

From 10^th^ March 2020, UHB included a structured COVID clerking sheet to capture COVID specific information in suspected cases. In Wave 1, 83.4% of swab confirmed COVID patients were suspected of having COVID at first presentation. In Wave 2, 64.3% of swab confirmed COVID patients were suspected of having COVID at first presentation.

Of those who were suspected of having COVID at initial presentation, presenting symptoms were collated. Apart from delirium (less prevalent), headache and chest pain (more prevalent), there were no differences between wave 1 and wave 2. These included (with Wave 1 data preceding wave 2 data) breathlessness (72.1% vs. 74.3%, p = 0.475); delirium (9.8% vs. 3.4%, p < 0.001), cough (68.4% vs. 71.8%, p = 0.280), sputum (10.4% vs. 13.8%, p = 0.122), headache (4.6% vs. 9.1%, p = 0.006), chest pain (4.3% vs. 11.9%, p < 0.001), fever (56.8% vs. 59.9%, p = 0.357), new diarrhoea or vomiting (7.3% vs. 11.0%, p = 0.053) and malaise (25.5% vs. 30.4% p = 0.667).

There was no difference in the median duration of symptoms prior to presentation (wave 1; 6 days (IQR 3 – 9 days) vs. wave 2, 6 days (IQR 3 – 8 days), p = 0.750).

### Disease severity, treatments and outcomes

Severity on initial presentation was available in 1494 patients across both waves. More people presented with mild disease in wave 2 compared to wave 1. In Wave 1, 57.7% were categorised as “mild”, 22.5% as “moderate” and 19.8% as having “severe” respiratory disease caused by COVID-19. In wave 2, 65.0% were categorised as having “mild” disease, 15.8% as “moderate” and 19.1% as “severe” disease, p =0.003. See figure 2. Of note, people who are South Asian and those over 85 were significantly more likely to have been admitted with mild severity in wave 2 compared with Wave 1.

**Figure 2.**
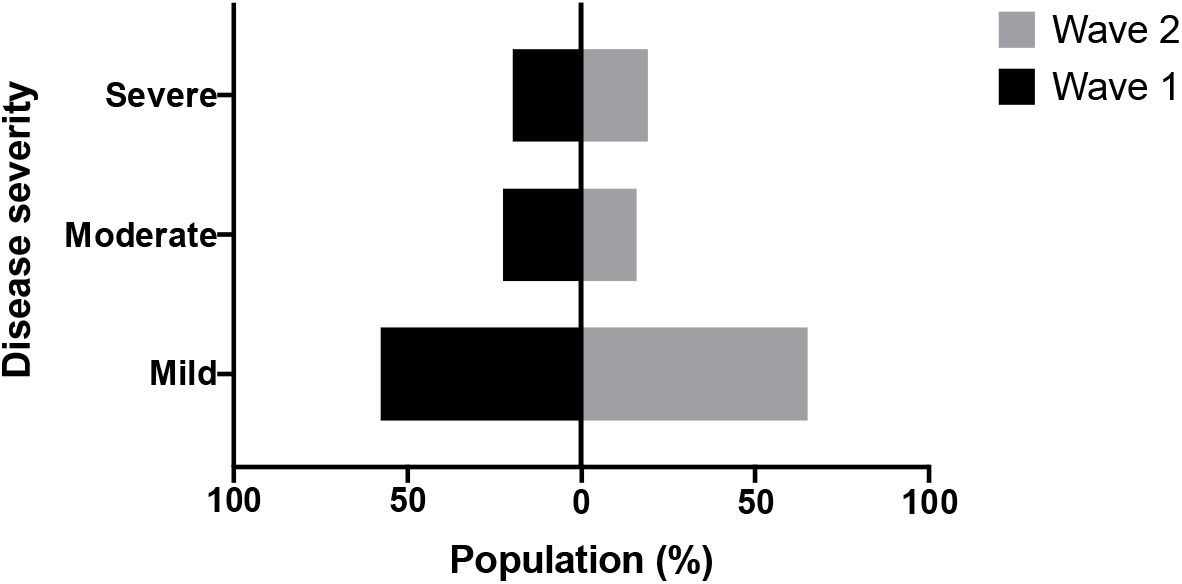
The severity of COVID-19 infection on initial presentation to hospital. **Legend**. The severity of respiratory disease on initial presentation to hospital. Data was available for 1494 patients, including 982 in wave 1 and 637 during wave 2 within the dates as given.

Prescribing data was available on 35% of the cohort in wave 1 and 33% of the cohort in wave 2 (those admitted to QEHB). See table 2. There was an increase in the percentage of patients receiving dexamethasone and remdesivir, in keeping with emerging evidence of efficacy(7, 25). There was a significant reduction in the prescription of broad-spectrum antibiotics, although the percentage of COVID-19 patients prescribed antibiotics remained high.

**Table 2.**
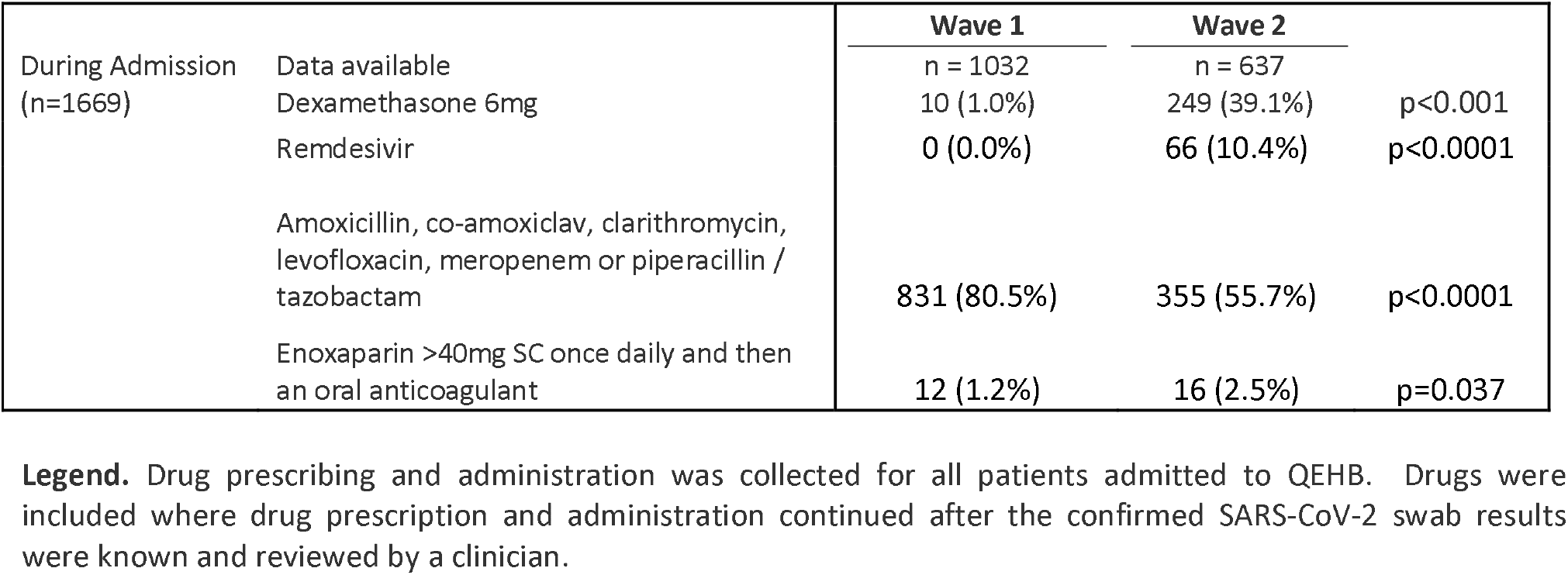
Drugs prescribed and administered for confirmed COVID-19 infections

At the time of study closure (14^th^ Dec 2020), no patient remained an in-patient from wave 1. The median length of stay in those discharged alive was 8 days (IQR 3 – 16 days) in wave 1. At the time of study closure (14^th^ Dec 2020), 104 patients remained as in-patients from Wave 2 (5.5%). Up until the censor date, patients has shown no difference in length of stay in those discharged alive (median 8 days (IQR 2 – 16).

There was no difference in the percentage of patients admitted to intensive care, with 356 (12.1%) of admitted patients in wave 1 and 232 (12.2%) of admitted patients during wave 2 were cared for in intensive care (p = 0.922). There was a reduction in mortality from COVID-19 in wave 2. 955 (32.3%) patients admitted during Wave 1 died within 28 days of a positive swab collection and 347 (18.2%) died in wave 2.

### Comparisons of mortality and intensive care admissions, accounting for age, ethnicity and co-morbidities

The whole dataset was matched on age (with 10 cut points), ethnicity (grouping Mixed, Chinese, and any Other Ethnicity together), and co-morbidity counts. 1512 (51.3%) of wave 1 data was matched with 1366 (71.6%) of the Wave 2 data. After matching for these factors, there remained a significant difference between the waves in terms of deaths within hospital where the odds of dying was approximately two-thirds less that of wave 1, and deaths within 28 days of a positive swab where the odds of dying was less than half of that in wave 1 (OR: 0.46 (0.38-0.55)), however there were no differences in the odds of admission to intensive care. See Table 3.

**Table 3.** The impact of Wave 1 versus Wave 2.

|  | Wave 1 | Wave 2 | p-value | Odds ratio (95% CI) * |
| --- | --- | --- | --- | --- |
| n | 1512 | 1366 |  |  |
| Admitted to ICU | 183 (12.1%) | 181 (13.3%) | $p=0.355$ | 1.11 (0.89-1.38) |
| Deaths Within hospital | 363 (24.0%) | 140 (10.2%) | $p<0.001$ | 0.36 (0.29-0.44) |
| Deaths within 28 days of swab being taken | 423 (28.0%) | 207 (15.2%) | $p<0.001$ | 0.46 (0.38-0.55) |

Legend: Patients were matched by age, ethnicity and co-morbidity counts and odds ratio of admission to intensive care or death (in hospital or within 28 days) was assessed. \*Reference group is Wave 1.

## Discussion

This paper to describes changes to the demography and outcomes of patients admitted to hospital over the course of the pandemic. In general, the study reports a reduction in the severity of infection with improved mortality rates in wave 2 compared to wave 1 in this hospitalised cohort. This is seen even when patient age, ethnicity and co-morbidities were accounted for (and wave 2 included a younger and more healthy population). However, a mortality rate of 18.2% is still extremely high for any condition. Further, the reduction in mortality did not equate to a reduction in the need for intensive care support, with approximately 12% of adults admitted with COVID-19 still requiring the highest level of supportive care, which is an important consideration for social policy and service planning.

Although patients admitted during wave 2 were generally more healthy than wave 1, there was still a high burden of co-morbidity and obesity, highlighting the continuing importance of these risk factors on patient outcomes(26, 27). Social deprivation was associated with hospital admission, with 54% of admissions in wave 2 coming from postcodes associated with the poorest IMD quintile. This is higher than would be expected for the local population, given that 40% of the Birmingham population and UHB catchment area live within this IMD quintile(28) but is in keeping with other reported studies(29).

The reasons for differences in the demography of wave 2 patients may reflect the impact of social restrictions which were widespread and common in Wave 2 including mask wearing in shops, the shielding of the elderly and more vulnerable.

Mortality was reduced, even when age, ethnicity and co-morbidities were accounted for, and the reasons for this are unclear. In part, this may reflect improved clinical management of these patients, including the use of treatments with proven efficacy. The reduction in broad-spectrum antibiotic prescribing may also reflect clinician awareness of the relatively low level of secondary bacterial infections in COVID-19(10), although the use of antibiotics was still high and the study did not assess reasons for clinician prescribing.

In the current study, less than two thirds of admitted wave 2 patients with confirmed COVID-19 were suspected as having COVID-19 at first presentation. It has been suggested that 12.5% of COVID cases within hospital were hospital acquired(30). The current study highlights the importance of routine screening for all patients, irrespective of presenting symptom.

It is reassuring that there were only 4 repeat admissions with a second confirmed hospitalised case of COVID-19, suggesting that although re-infection has been reported(31), it remains uncommon (at least prior to the dominance of new variants).

This study has a number of strengths. This includes the study of a highly curated and complete data set, without the inherent issues of significant under-coding seen with morbidity and ethnicity data when using a secondary care dataset. The regional approach within a non-transient population allows for the natural history of hospitalised COVID-19 admitted patients to be described in the context of changing national and regional legislation and social policy. It provides a strong foundation from which to assess the impact of new variants of COVID and the national vaccine programme.

There are limitations to this study. First, 5.5% of patients from wave 2 remained in hospital at the time of the data lock (albeit after 28 days from their positive swab) and more patients have been admitted, so our findings will evolve. Second, this study does not include confirmation of genetic variation in the SARS-CoV-2 virus for all patients, but results are based on screening data from the UK. Third, this study focuses on hospitalised care and we are therefore unable to comment upon the natural history of COVID-19 prior to admission to secondary care or in those without hospital admission.

In summary, there are differences in the demography, co-morbid disease burden, COVID-severity and outcomes in patients hospitalised with COVID-19 comparing wave 1 and wave 2 timelines, with wave 2 generally associated with better outcomes. The reduction in mortality was seen even after accounting for age, ethnicity and co-morbidity. However, mortality remained high and intensive care admissions were not reduced, highlighting the continuing impact of SARS-CoV-2 on our population and healthcare systems. This dataset will provide a robust means to compare the severity of hospitalised illness associated with any new variant of COVID-19, and the potential impact of vaccination, assisting with service planning during this pandemic.

## Data Availability

Data Access:  Data from this study is available from PIONEER, the Health Data Hub in Acute care, in accordance with Hub processes.  See www.pioneerdatahub.co.uk and contact for more details.

## Acknowledgements

This work was supported by PIONEER, the Health Data Research Hub in acute care, and the Better Care Programme, both funded by Health Data Research UK. This work uses data provided by patients and collected by the NHS as part of their care and support. We would like to acknowledge the contribution of all staff, key workers, patients and the community who have supported our hospitals and the wider NHS at this time.

## Author contributions

C.A, V.K, V.R-K, D.P assisted with clinical data insights, F.E, P.N and S.G analysed the data. C.A, S.G, S.B and E.S designed the study and wrote the manuscript. All authors approved the final manuscript.

## Data Access

Data from this study is available from PIONEER, the Health Data Hub in Acute care, in accordance with Hub processes. See www.pioneerdatahub.co.uk and contact for more details.

